# The impact of seasonal respiratory virus transmission on syndromic surveillance for COVID-19 in Ontario, Canada

**DOI:** 10.1101/2020.12.02.20242735

**Authors:** Arjuna Maharaj, Jennifer Parker, Jessica P. Hopkins, Effie Gournis, Isaac I. Bogoch, Benjamin Rader, Christina M. Astley, Noah Ivers, Jared B. Hawkins, Ashleigh R. Tuite, David N. Fisman, John S. Brownstein, Lauren Lapointe-Shaw

## Abstract

**Background:** Syndromic surveillance systems for COVID-19 are being increasingly used to track and predict outbreaks of confirmed cases. Seasonal circulating respiratory viruses share syndromic overlap with COVID-19, and it is unknown how they will impact the performance of syndromic surveillance tools. Here we investigated the role of non-SARS-CoV-2 respiratory virus test positivity on COVID-19 two independent syndromic surveillance systems in Ontario, Canada.

**Methods:** We compared the weekly number of reported COVID-19 cases reported in the province of Ontario against two syndromic surveillance metrics: 1) the proportion of respondents with a self-reported COVID-like illness (CLI) from COVID Near You (CNY) and 2) the proportion of emergency department visits for upper respiratory conditions from the Acute Care Enhanced Surveillance (ACES) system. Separately, we plotted the percent positivity for other seasonal respiratory viruses over the same time period and reported Pearson’s correlation coefficients before and after the uncoupling of syndromic tools to COVID-19 cases.

**Results:** There were strong positive correlations of both CLI and ED visits for upper respiratory causes with COVID-19 cases up to and including a rise in entero/rhinovirus (r = 0.86 and 0.87, respectively). There was a strong negative correlation of both CLI and ED visits for upper respiratory causes with COVID-19 cases (r = −0.85 and −0.91, respectively) during a fall in entero/rhinovirus.

**Interpretation:** Two methods of syndromic surveillance showed strong positive correlations with COVID-19 confirmed case counts before and during a rise in circulating entero/rhinovirus. However, as positivity for enterovirus/rhinovirus fell in late September 2020, syndromic signals became uncoupled from COVID-19 cases and instead tracked the fall in entero/rhinovirus. This finding provides proof-of-principle that regional transmission of seasonal respiratory viruses may complicate the interpretation of COVID-19 surveillance data. It is imperative that surveillance systems incorporate other respiratory virus testing data in order to more accurately track and forecast COVID-19 disease activity.

---

Emerging evidence is showing that syndromic surveillance systems for COVID-19 can be used to predict outbreaks of confirmed cases with high spatial and temporal resolution.^1–3^ These methods can be used as an early warning system to guide regional public health policy decisions. Tools include passive methods (e.g. tracking healthcare encounters), as well as more active participatory surveillance, whereby individuals self-report symptoms using phone or internet.^2,3^ Currently, it is unknown how seasonal circulating respiratory viruses impact the performance of COVID-19 surveillance tools, although their symptomatic overlap makes this a theoretical concern.^4^ We investigated the role of non-SARS-CoV-2 respiratory virus test positivity on two independent COVID-19 syndromic surveillance systems in Ontario, Canada.

## METHODS

Our data sources were: 1) self-reported symptoms from COVID Near You (CNY), 2) emergency department (ED) visits for upper respiratory conditions from the Acute Care Enhanced Surveillance (ACES) system, 3) provincial laboratory-confirmed COVID-19 case counts and 4) percent positivity for other respiratory viruses reported by Public Health Ontario, from April 20^th^, 2020 to November 1^st^. COVID-19-like illness (CLI) from CNY was defined using the CDC Surveillance Case Definition for COVID-19.^5^ ACES uses machine learning algorithms to categorize ED visit chief complaints into clinical syndromes.^6^ See supplemental methods for full description on data sources and syndromic definitions.

We compared the weekly (by International Organization for Standardization (ISO) date week number of reported COVID-19 cases against the proportion of CNY respondents with CLI and the proportion of all emergency department visits for an upper-respiratory condition. Separately, we plotted the percent positivity for other respiratory viruses over the same time period. We reported Pearson’s correlation coefficients before and after the uncoupling of syndromic tools to COVID-19 cases. The data were analyzed using R version 4.0.1 in the RStudio software environment, version 1.1.463 (RStudio Inc., Boston, MA).

The study was approved by the Research Ethics Board of the University of Toronto and a waiver of informed consent was granted because the data were collected for public health surveillance purposes.

## RESULTS

There were strong positive correlations of both CLI and ED visits for upper respiratory causes with COVID-19 cases up to and including week 40 (r = 0.86 and 0.87, respectively). Subsequently, from week 41–45 there was a strong negative correlation of both CLI and ED visits for upper respiratory causes with COVID-19 cases (r = −0.85 and −0.91, respectively). There was also an observed rise in enterovirus/rhinovirus percent positivity in Ontario from weeks 35 to week 39, to a peak of 22.8% in week 39, and subsequent fall in weeks 39 – 44 (figure 1B).

**Figure 1.**
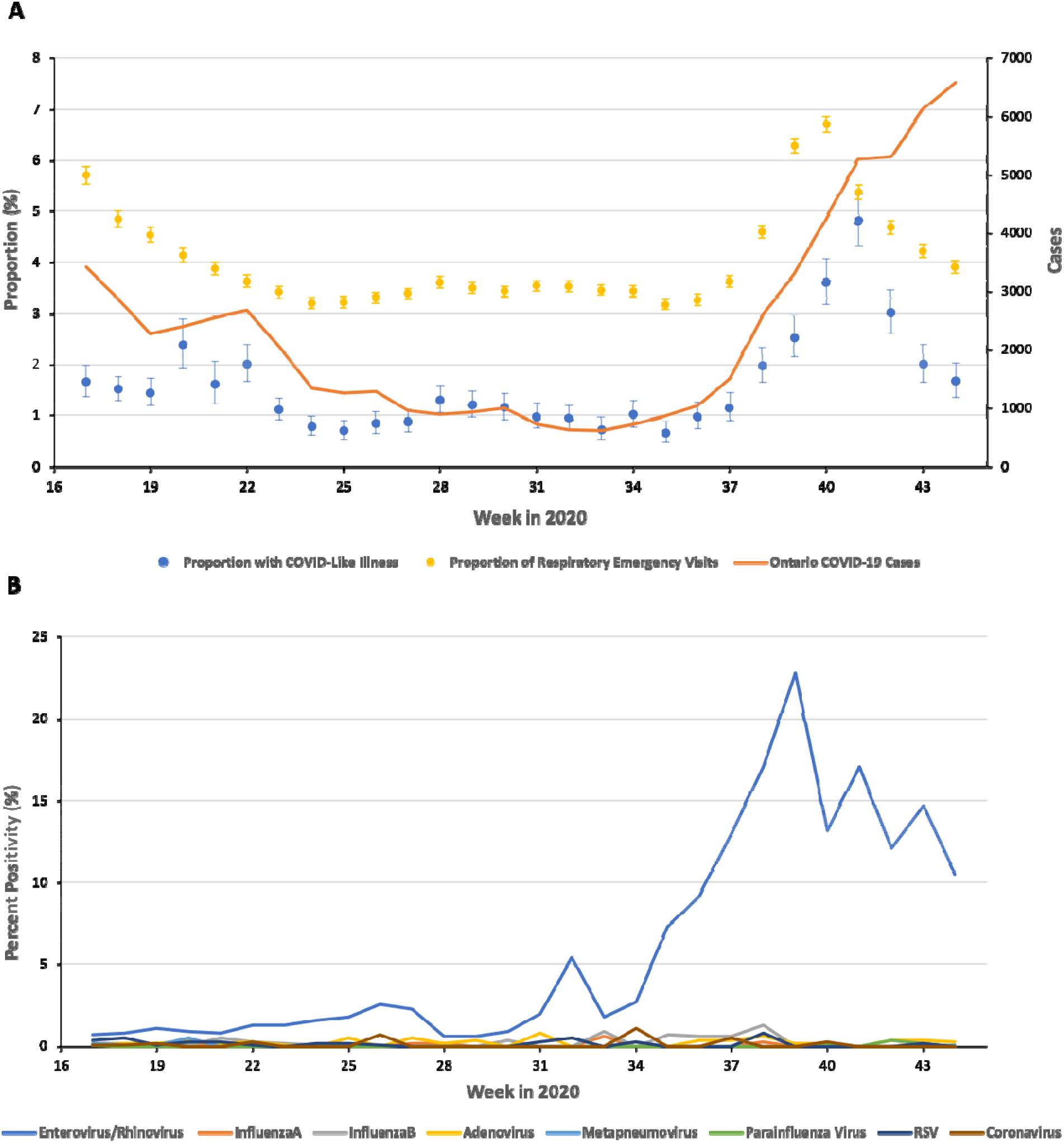
(A) Weekly syndromic surveillance signals and weekly cases in Ontario. (B) Weekly percent positivity of respiratory viruses in Ontario. Dashed line represents onset of rise of Rhinovirus/Enterovirus at week 34. Error bars represent Clopper-Pearson interval

## DISCUSSION

Two methods of syndromic surveillance showed strong positive correlations with COVID-19 confirmed case counts before and during a rise in circulating entero/rhinovirus. However, as positivity for enterovirus/rhinovirus fell in late September 2020, syndromic signals became uncoupled from COVID-19 cases. It is now apparent that although these seemed to be tracking closely in weeks 34-40, the rise in syndromic cases in this period reflected rapidly rising entero/rhinovirus disease activity rather than COVID-19. Even milder, seasonal respiratory viruses such as rhinoviruses have considerable syndromic overlap with COVID-19^4^. The absence of an envelope may explain why rhinoviruses tend to be more robust in diverse environments, and why they have spread so easily despite current public health measures. This finding provides proof-of-principle that regional transmission of seasonal respiratory viruses may complicate the interpretation of COVID-19 surveillance data. It is imperative that surveillance systems incorporate other respiratory virus testing data in order to more accurately track and forecast COVID-19 disease activity.

## Data Availability

The data that support the findings of this study are available from the corresponding author, LLS, upon reasonable request.

## ACKNOWLEDGEMENT

ACES is funded by the Ministry of Health and administrative support is provided by Kingston, Frontenac and Lennox & Addington Public Health (for more information, see www.kflaphi.ca).

IIB has consulted to BlueDot, a social benefit corporation that tracks the spread of emerging infectious diseases

Data for this study were obtained from the Ontario Ministry of Health and Long-term Care as part of the province’s emergency modeling table.

## Notes

### Funding Statement

No external funding was received for this work

### Summary of Updates

Author affiliations and order updated

## REFERENCES

1. Desjardins MR. Syndromic surveillance of COVID-19 using crowdsourced data. Lancet Reg Heal - West Pacific. Published online 2020. doi:10.1016/j.lanwpc.2020.100024

2. Chan AT, Brownstein JS. Putting the public back in public health - Surveying symptoms of Covid-19. N Engl J Med. Published online 2020. doi:10.1056/NEJMp2016259

3. Nomura S, Yoneoka D, Shi S, et al. An assessment of self-reported COVID-19 related symptoms of 227,898 users of a social networking service in Japan: Has the regional risk changed after the declaration of the state of emergency? Lancet Reg Heal - West Pacific. Published online 2020. doi:10.1016/j.lanwpc.2020.100011

4. Jiang F, Deng L, Zhang L, Cai Y, Cheung CW, Xia Z. Review of the Clinical Characteristics of Coronavirus Disease 2019 (COVID-19). J Gen Intern Med. Published online 2020. doi:10.1007/s11606-020-05762-w

5. Centers for Disease Control and Prevention. Coronavirus Disease 2019 (COVID-19) 2020 Interim Case Definition. Published 2020. https://wwwn.cdc.gov/nndss/conditions/coronavirus-disease-2019-covid-19/case-definition/2020/

6. KFL&A Public Health. Acute Care Enhanced Surveillance (ACES). Published 2019. Accessed November 1, 2020. http://www.kflaphi.ca/acute-care-enhanced-surveillance/#

