## Supplemental Methods for "The impact of seasonal respiratory virus transmission on syndromic surveillance for COVID-19 in Ontario, Canada"

1.) Ontario Respiratory Virus Data

Data on the percent positivity of non-SARS-CoV2 respiratory pathogens were obtained from the Ontario Respiratory Pathogen Bulletin (ORPB). This provides a weekly summary of the laboratory-confirmed percent positivity of 8 common viruses in Ontario. This data is submitted to the Public Health Agency of Canada from 16 participating laboratories in Ontario, including 11 Public Health Ontario Laboratories and five hospital-based laboratories. Data was extracted on November 1^st^, 2020.

2.) COVID Near You

COVID Near You (covidnearyou.org) is a web-based participatory health surveillance tool created by infectious disease epidemiologists at Boston Children’s Hospital and launched March 2020. This team also created Flu Near You (flunearyou.org), a similar tool for influenza symptoms, which has been validated against clinical data sources and applied to predict influenza trends. Participants are asked to report on present symptoms, date of symptom onset, demographic information, healthcare encounters, testing, and results. We included responses with a self-reported postal code originating from Ontario, Canada, between April 20th, 2020 (week 17) and November 1^st^, 2020 (week 44).

Symptoms of possible COVID-19 were defined using the CDC Surveillance Case Definition for COVID-19 from the National Notifiable Diseases Surveillance System (NNDSS) approved August 5^th^, 2020 to develop a COVID-like illness (CLI) metric. The criteria for a CLI required at least two of the following symptoms: fever (measured or subjective), chills, rigors, myalgia, headache, sore throat or at least one of the following symptoms: cough, shortness of breath, difficulty breathing, new olfactory disorder, or new taste disorder.

3.) Acute Care Enhanced Surveillance

ACES receives real or near-real time patient registration data from over 95% of Ontario’s acute care hospitals. ACES uses machine learning algorithms to categorize ED visits into syndromes validated to medical diagnoses using chief complaints from triage notes. The ACES Respiratory syndrome captures descriptions of symptoms including words or phrases describing respiratory ailments such as “cough”, “sore throat”, “upper respiratory tract infection”, and “sinus infection”. ACES Respiratory syndrome is validated against daily aggregates of the following ICD-10 medical diagnoses reported by National Ambulatory Care Reporting System:

J00 (acute nasophyngitis), R05 (cough), J01 (Acute Sinusitis), J02 (Acute Pharyngitis), J03 (Acute tonsillitis), J04 (Acute laryngitis and tracheitis), J05 (Acute epiglottitis), J06 (Acute upper respiratory infections of multiple and unspecified sites), J22 (Unspecified acute lower respiratory infection), J31 (Chronic Rhinitis), J32 (Chronic Sinusitis), J37 (Chronic laryngitis), J39 (Other diseases of upper respiratory tract)

4.) COVID-19 Case Counts in Ontario

Case and Contact Management (CCM) Plus data system has been implemented in Ontario to record COVID-19 case information. Each of Ontario’s 34 public health units is responsible for local COVID-19 case investigation and entry of case information into CCM Plus. Ontario’s case definition for a confirmed case of COVID-19 has evolved based on science and available testing methods, but generally requires a positive laboratory test using a validated nucleic acid amplification test, including real-time PCR and nucleic acid sequencing. Confirmed COVID-19 case counts were obtained from the CCM Plus data system on November 1^st^, 2020 for the time period between April 20^th^ and November 1^st^, 2020.
